# Self-Other Voice Discrimination Task: A - Neuropsychological Tool For Clinical Assessment of Self-Related Deficits

**DOI:** 10.1101/2024.03.04.24303420

**Authors:** Philippe Voruz, Pavo Orepic, Selim Yahia Coll, Julien Haemmerli, Olaf Blanke, Julie Anne Péron, Karl Schaller, Giannina Rita Iannotti

**Author notes:** **Corresponding author:** Philippe Voruz, Faculty of Psychology and Educational Sciences, 40 bd du Pont d’Arve, 1205 Geneva, Switzerland. Equal contribution (Karl Schaller, Giannina Rita Iannotti).

## Abstract

**Background:** Deficits in self are commonly described through different neuro-pathologies, based on clinical evaluations and experimental paradigms. However, currently available approaches lack appropriate clinical validation, making objective evaluation and discrimination of self-related deficits challenging.

**Methods:** We applied a statistical standardized method to assess the clinical discriminatory role of a Self-Other Voice Discrimination (SOVD) task. This task, validated experimentally as a marker for self-related deficits, was administered to 17 patients eligible for neurosurgery due to focal hemispheric brain tumors or epileptic lesions.

**Results:** The clinical discriminatory capacity of the SOVD task was evident in three patients who exhibited impairments for self-voice perception that could not be predicted by other neuropsychological performances. Impairments in other-voice perception were linked to inhibitory neuropsychological alterations, suggesting a potential association with executive deficits in voice recognition.

**Conclusions:** This exploratory study highlights the clinical discriminatory potential of the SOVD task and suggests that it could complement the standard neuropsychological assessment, paving the way for enhanced diagnoses and tailored treatments for self-related deficits.

## 1. Introduction

The assessment self-related deficits is of great importance in clinical care whether in the context of acquired neurological pathologies or before/following interventions (e.g., neurosurgical) ^1^. However, the current understanding and diagnosis of self-related deficits are constrained by the lack of clinical tests that are psychometrically validated as standard neuropsychological tools. Indeed, the standard neuropsychological assessments of self-related deficits rely on the clinician’s subjective evaluation, questionnaires or a comparison between objective performance and the patient’s subjective evaluation ^2 3^. In this context, an objective evaluation derived from experimental paradigms investigating the self, based on psychometrical analysis (e.g., quantitative values) adapted to clinical practice (i.e. using validated clinical cut-offs) is of utmost importance to detect potential self-related deficits. Here, we explore the potential use of Self-Other Voice Discrimination (SOVD) task for this purpose, as it is an experimentally validated marker for self-related deficits ^4-6^.

Given the intimate role self-voice plays in our identity, and that self-voice misperception has been related to psychotic symptoms such as auditory hallucinations ^7^, a specialized clinical tool integrating self-voice perception could be promising for detecting self-related deficits. To date, some studies successfully attempted the objectivation (e.g., quantification) of voice recognition abilities within clinical neuropsychological examinations, by indicating specific brain regions and mechanisms which, if altered, can lead to a voice recognition disorder known as phonagnosia ^8^. However, these studies did not integrate self-voice. Recently, we introduced a SOVD task, highlighting perceptual and neural mechanisms underlying self-voice perception in healthy controls (HC) ^5 6^, and demonstrating its clinical potential in identifying personality alterations after neurosurgical intervention ^5^. Similarly, others have used self-voice tasks to delineate different types of brain damage with respect to self ^9^. However, previous clinical studies do not provide any clinical value/cut-off for categorizing a patient’s performance as normal or deficient, in comparison with expected performance in HC. In other words, these studies lack of a dedicated analysis allowing to categorize the individual performance and assess the prevalence of self-related deficits.

In line with the methodology followed to characterize the phonagnosia, the aim of the current exploratory study was to evaluate the discriminatory potential of the SOVD task, in terms of standardized psychometric measures with analyses at individual level. To that purpose, 17 neurosurgical patients, prior to their neurosurgical intervention, were enrolled in a SOVD task and their performance was analyzed in comparison to standardized data obtained from a comparable group of HC. We hypothesized that the SOVD task would discriminate self-related alteration based on patients’ performance.

## 2. Methodology

### 2.1. Participants

We included 17 patients (5 females; M_age_= 46.17; SD_age_ = 14.94; 10 right-hemisphere lesions) at the University Hospitals of Geneva (HUG) suffering from a focal hemispheric brain tumor (15 patients) or having a focal epileptic lesion (2 patients), who were candidates for neurosurgery (see Table 1). Based on power analysis carried out on a study that evaluated self-voice in patients with stroke ^9^, a sample of 15 patients was considered sufficient to achieve the (1 - β) of 90% and a risk of Type I error (α) of 0.05, in a two-sided hypothesis.

**Table 1.** Sociodemographic, clinical and neuropsychological characteristics of patients.

| Patient | Clinical characteristics |  |  | Memory |  |  |  | Executive |  |  |  | Instrumental |  |  |  |  | Attention |
| --- | --- | --- | --- | --- | --- | --- | --- | --- | --- | --- | --- | --- | --- | --- | --- | --- | --- |
|  | Affected hemisphere | Type of lesion | Localisation | verbal episodic | V-S episodic | verbal short-term | V-S short-term | Program-ming | Mental flexibility | Inhibition | Verbal Working memory | V-S memory | Verbal fluency | Perception | Language prod. | Language comp. | Attention |
| 1 | R | Oligodendroglioma OMS 2 | Insula and fronto-opercular pars opercularis | DEF | NOR | NOR | NOR | DEF | DEF | DEF | DEF | DEF | DEF | NOR | NOR | NOR | DEF |
| 2 | L | Glioma | Superior frontal gyrus and cingulate gyrus | NOR | NOR | NOR | NOR | NOR | NOR | NOR | NOR | NOR | NOR | NOR | NOR | NOR | NOR |
| 3 | R | Astrocytoma grade II | Temporo-mesial and insular | DEF | NOR | NOR | NOR | DEF | NOR | NOR | NOR | NOR | NOR | NOR | NOR | NOR | DEF |
| 4 | R | Astrocytoma grade II | Middle frontal gyrus | NOR | NOR | NOR | NOR | NOR | NOR | NOR | NOR | NOR | NOR | NOR | NOR | NOR | NOR |
| 5 | L | Glioblastoma | Temporo-occipital junction | DEF | NOR | NOR | NOR | NOR | DEF | NOR | NOR | NOR | NOR | NOR | DEF | NOR | DEF |
| 6 | R | Glioblastoma | Insula | DEF | NOR | NOR | NOR | NOR | NOR | NOR | NOR | NOR | NOR | NOR | NOR | NOR | NOR |
| 7 | R | Glioblastoma | Hippocampus and temporo-parietal | NOR | NOR | NOR | NOR | NOR | NOR | NOR | DEF | NOR | DEF | DEF | NOR | NOR | DEF |
| 8 | L | Astrocytoma grade II | Superior frontal and anterior gyrus | NOR | NOR | NOR | NOR | NOR | NOR | DEF | NOR | NOR | NOR | NOR | NOR | NOR | NOR |
| 9 | R | Glioblastoma | fronto-opercular pars opercularis and triangularis | DEF | DEF | NOR | NOR | NOR | DEF | NOR | NOR | NOR | DEF | NOR | NOR | NOR | DEF |
| 10 | R | Meningioma | Frontal | NOR | NOR | NOR | NOR | DEF | NOR | NOR | DEF | NOR | NOR | NOR | DEF | NOR | NOR |
| 11 | L | Ganglioglioma | Superior temporal gyrus | NOR | NOR | NOR | NOR | NOR | NOR | NOR | NOR | NOR | DEF | NOR | DEF | NOR | NOR |
| 12 | R | Astrocytoma grade II | Hippocampus | NOR | NOR | NOR | NOR | NOR | NOR | DEF | NOR | NOR | NOR | NOR | NOR | NOR | DEF |
| 13 | L | Epilepsy | Fronto-temporal | NOR | NOR | NOR | NOR | NOR | NOR | NOR | NOR | NOR | NOR | NOR | NOR | NOR | NOR |
| 14 | R | Astrocytoma grade II | Frontal | DEF | NOR | NOR | NOR | DEF | NOR | DEF | DEF | NOR | DEF | NOR | DEF | NOR | NOR |
| 15 | L | Epilepsy | Fronto-temporal | DEF | DEF | NOR | NOR | DEF | DEF | NOR | NOR | NOR | NOR | NOR | DEF | NOR | DEF |
| 16 | L | Metastasis | Fronto-opercular | DEF | NOR | NOR | ND | DEF | NOR | NOR | NOR | ND | DEF | DEF | NOR | NOR | DEF |
| 17 | L | Glioblastoma | Parietal lobe | ND | ND | ND | ND | ND | ND | ND | ND | ND | ND | ND | ND | ND | ND |

**Legend.** DEF: Deficit for the neuropsychological function evaluated (< -1.00 Z-scores; < Percentile 16; < 40 T-score); F: Female; L: Left; M: Male; NOR: Normal [with conservative threshold] neuropsychological performance; R: Right; V-S: Visuo-spatial.

To define clinical psychometric values, we extracted data from 17 HC (9 females; M_age_ = 37.29; SD_age_ = 15.58) from a previous study ^4^.

All participants reported no hearing deficits. Moreover, participants’ clinical history was evaluated and no psychiatric condition anterior to the evaluation was objected. All participants gave their informed consent prior to their participation and received monetary compensation (CHF 20/h).

The study was conducted in accordance with the Declaration of Helsinki and received approval from the national Commission Cantonal d’Ethique de la Recherche de Geneva (CCER, code approved protocol PB_2016-01635).

### 2.2. Neuropsychological assessment

Retrospectively, we extracted the neuropsychological examinations performed by board-certified neuropsychologists in the context of the pre-surgical clinical assessment for all patients except one (the neuropsychological assessment could not be retrieved). Neuropsychological evaluations were not standardized for the type of test. As an example, different tests were performed for verbal episodic memory (e.g., MEM-III ^10^; RL/RI16 ^11^). However, for all evaluated neuropsychological functions (memory; executive; attentional; instrumental), psychometric data allowed to determine the presence or absence of cognitive deficits (see Table 1). Therefore, based on validated neuropsychological methods to calculate the prevalence of deficits in cognitive measures ^12^, and according to the guidelines of the Swiss Association of Neuropsychology ^13 14^, a non-conservative cut-off (< -1.00 Z-scores; < Percentile 16; < 40 T-score) was applied.

### 2.3. Self-Other voice discrimination task (SOVD)

Each patient participated in the SOVD task, following the procedure described in our previous works on HC ^4^. Concisely, each patient was asked to discriminate between Self and Other’s voice from a continuum of randomly presented voice-morphs, featuring varying levels of patient’s voice presence: 15%, 30%, 45%, 55%, 70%, and 85%.

### 2.4. Statistical analysis

First, patients’ performances on the SOVD task was measured in terms of score of accuracy to detect self/other voice-morphs. Global score; Self-voice score; Other-voice score were extracted and Z-scored, based on HC performance as follows:

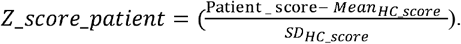

This standardization allowed to control for age and gender effects. The patient’s z-scores were then categorized (deficit/non-deficit) according to the conservative clinical cut-offs established by the Swiss Association of Neuropsychology (deficit: < - 1.60 Z-score).

In order to exclude the potential effects of confounding factors, 6 distinct models of backward stepwise multiple regression were carried out: *i) for neuropsychological predictors*: between the standardized SOVD z-scores (i.e..: Self-voice; Other-voice; Global) and neuropsychological performances; ii) *for other clinical predictors*: between the standardized SOVD z-scores (i.e., Self-voice; Other-voice; Global;) and secondary clinical variables (lateralization of lesions; type of lesion).

## 3. Results

### 3.1. Prevalence of deficits

No significant differences between patients and HC were found in terms of sociodemographic data.

Analysis of standardized scores revealed that 5 patients (29.41% of the sample) presented deficits in performance for the Global score on the SOVD task and 12 patients (70.59% of the sample) displayed comparable performances (for all SOVD measures) to the HC group. By considering the specific conditions (Self-voice and Other-voice), 2 out of the 5 patients (11.76% of the global sample) presented an impaired score for the recognition of both the Self- and Other-voice, suggesting a general deficit in voice discrimination (e.g., phonagnosia). Interestingly, 3 patients (17.65% of the global sample) had difficulties specific to Self-voice recognition, in the absence of other-voice or general voice recognition deficits, suggesting a potential self-related deficit.

**Figure 1.**
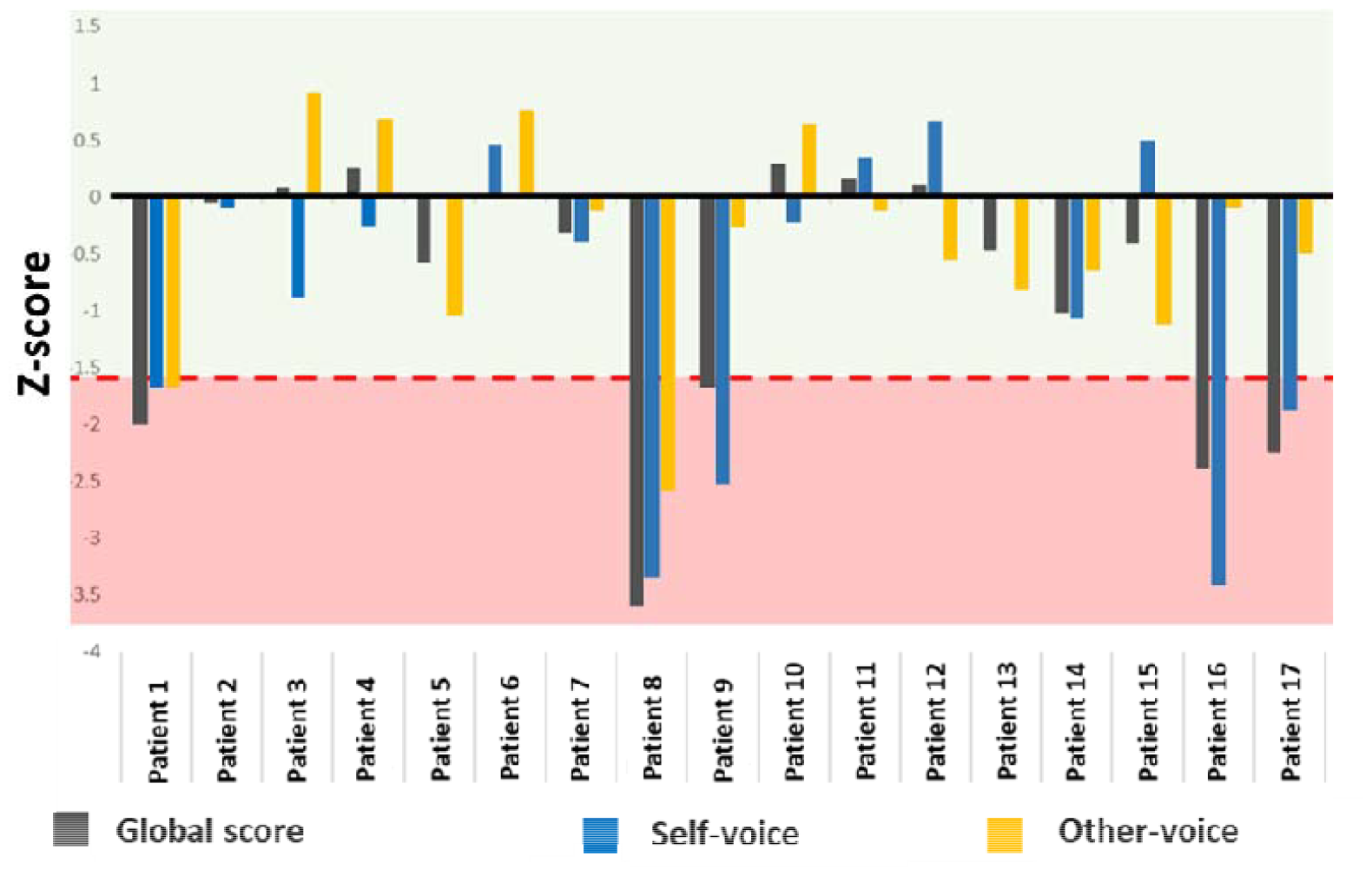
Z-standardized scores for the self-other voice discrimination (SOVD) task scores. Based on Swiss Association of Neuropsychology (ASNP) guidelines, a threshold of -1.60 Z-score (delimited by the red line) was set for the definition of impaired performance. impaired performances were observed for 5 patients for the global score (gray bars); 3/5 had a specific deficit of Self-voice recognition (Patients 9,16,17),

### 3.2. Confounding factors

For the evaluation of *neuropsychological predictors*, we observed significant results only for the Other-voice condition performance, which was associated with the presence/absence of inhibitory deficits (*t* = 2.87, *p* = .013). All other predictors were non-significant (*all p’s* > .051).

For the evaluation of *other clinical predictors* (i.e. hemisphere of lesion and type of lesion), analysis did not reveal significant results (*all p’s* > .08).

## 4. Discussion

Here we conducted an exploratory analysis on a cohort of neurosurgical patients who were enrolled in a SOVD task, in the context of a previous research project. The aim was to evaluate, through standardized statistical analyses commonly used in neuropsychological psychometrics, the clinical discriminatory potential of the SOVD task for the objectification of self-related voice deficit. Although the majority of the patients considered in this work were comparable to HC in terms of task performance, the SOVD task allowed to discriminate distinct clinical phenomena. Indeed, among patients presenting impaired performance (5), it was possible to distinguish a subgroup of patients (2) likely affected by phonagnosia (impaired performance both for Self- and Other-voice discrimination), and a subgroup of patients (3) characterized by self-related deficits, manifested by impaired performances specific to the Self-voice recognition condition. These results reveal the potential of the SOVD task in targeting Self-voice impairments, independently from the presence of deficits in other neuropsychological functions assessed. In the case of recognition of the Other-voice condition, the presence/absence of inhibitory deficits was significantly associated with Other-voice trials, suggesting the involvement of executive processes in voice processing, as has already been demonstrated previously ^15^. Other clinical variables, such as hemispheric lateralization of the lesion, type of pathology, and the presence of neuropsychological deficits were not predictive of the SOVD performance, in particular for the Self-voice condition. This indicates that distinct self-related deficits, could not be accounted by these factors in our heterogeneous cohort. Although our results on the 3 patients with self-related deficits lead us to hypothesize the involvement of frontal and parietal regions (Table 1), in accordance with the literature ^1^, future work with homogeneous lesions location and type is needed to address lesion-specific effects on the SOVD task performance.

However, our current results underline the importance to integrate adapted experimental tools for assessing the sense of self in the clinical practice, by emphasizing the relevance of self-voice-based tasks. In addition, an objective assessment of self-related impairments would enhance the comprehension of patients’ symptomatology and could be useful for personalized neuropsychological rehabilitation. Importantly, considering its short duration (∼15 minutes), the SOVD task could find practical applicability in the neuropsychological routine, across various pathologies.

In the future a specifically dedicated psychometric validation on increased control and clinical groups would be considered to assess potential biases. In addition, the inclusion of standardized neuropsychological assessment for the evaluation of associated factors (e.g., effect of executive and attentional deficits) would be important to encompass the limitations inherent to the neuropsychological assessments available for our sample. Furthermore, although possible auditory impairments do not affect the SOVD performance (given the counterbalancing of conditions), it would be useful to integrate explicit analysis for auditory performance (e.g., pitch recognition). In addition, to better characterize the mechanisms inherent to voice-identity recognition, it would be important to develop the SOVD task and integrate vocal stimuli pronounced by self/familiar and unfamiliar voices. In conclusion, this work shows that the SOVD task could serve as a psychometrically validated tool in the clinical setting for an objective standardized neuropsychological assessment of self-related deficits.

## Data Availability

All data produced in the present study are available upon reasonable request to the authors

## Acknowledgement

We would like to thank the patients for contributing their time to this study.

